# A national consensus management pathway for Paediatric Inflammatory Multisystem Syndrome - Temporally associated with SARS-CoV-2 (PIMS-TS): The results of a national Delphi process

**DOI:** 10.1101/2020.07.17.20156075

**Authors:** Rachel Harwood, Benjamin Allin, Christine E Jones, Elizabeth Whittaker, Padmanabhan Ramnarayan, Athimalaipet Ramanan, Musa Kaleem, Robert Tulloh, Mark J Peters, Sarah Almond, Peter J Davis, Michael Levin, Saul N. Faust, Marian Knight, Simon Kenny, on behalf of the PIMS-TS National Consensus Management Study Group

## Abstract

**Objective:** To develop a consensus management pathway for children with Paediatric Inflammatory Multisystem Syndrome - Temporally associated with SARS-CoV-2 (PIMS-TS).

**Design:** A three-phase online Delphi process and virtual consensus meeting sought consensus over the investigation, management and research priorities from 98 multidisciplinary participants caring for children with PIMS-TS. 46 participants (47%) completed all three phases. Participants were grouped into three panels and scored each statement from 1 (disagree) to 9 (strongly agree). In phase two participants were shown their panel’s scores, and in phase three all panels’ scores.

Consensus agreement was defined as ≥70% of participants in each panel scoring the statement 7-9, and <15% scoring 1-3, and consensus disagreement was the opposite of this. Statements which achieved consensus in 2/3 panels were discussed at the consensus meeting, and when ≥70% participants agreed with the statement it achieved consensus.

**Results:** 255 statements were assessed, with ‘consensus agreement’ achieved for 111 (44%), ‘consensus disagreement’ for 29 (11%), and no consensus for 115 (45%). The 140 consensus statements were used to derive the consensus management pathway.

**Conclusions:** A national consensus pathway has been developed for children suspected of having the novel syndrome PIMS-TS in a timely, cost-efficient manner, in the midst of a global pandemic. Use of a rapid online Delphi process has made this consensus process possible. Future evidence will inform updates to this guidance, which in the interim provides a solid framework to support clinicians caring for children with PIMS-TS.

## Introduction

Since the first reports from London, UK, in late April 2020, many countries globally have reported children presenting severely unwell with features of significant inflammation temporally related to the COVID-19 pandemic. These include the United States of America(1), France(2, 3), Italy(4) and the United Kingdom(5, 6). Subsequently, parallels have been drawn between the presenting features of this syndrome and other known conditions, including complete, incomplete and atypical Kawasaki Disease (with or without coronary artery dilatation), toxic shock syndrome, viral sepsis and less commonly, Macrophage Activation Syndrome (MAS) or Haemophagocytic Lymphohistiocytosis (HLH). Preliminary case definitions of this novel inflammatory condition have been published by the Royal College of Paediatrics and Child Health (RCPCH)(7), the Centre for Disease Control(8) and the World Health Organisation(9). As these definitions are based on relatively small numbers of children seen, variation exists. For the purposes of this paper, which focuses on the opinions of UK clinicians, the RCPCH definition, which names the condition ‘*Paediatric Inflammatory Multisystem Syndrome - Temporally associated with SARS-CoV-2 (PIMS-TS)’* has been used.

It rapidly became apparent that there are many clinical uncertainties regarding this new disease syndrome. These include the prevalence, apparent differing clinical phenotypes, variable severity, the clinical course, and optimal management. To provide clarity to UK clinicians, NHS England led a process to develop national clinical management guidance through a rapid consensus exercise. The process also explored where equipoise exists for the planning of formal research trials including children with PIMS-TS. Given the status of PIMS-TS as a new syndrome, clinical consensus combined with experience in treating the initial cases was the starting point in the process of constructing a clinical guideline and defining key areas of research. The UK Randomised Evaluation of COVID-19 (RECOVERY) trial (https://www.recoverytrial.net/) steering committee made the trial protocol (including anti-inflammatory agents) available to children with COVID-19 and related inflammation prior to NHS England initiating the consensus process. Therefore, enrolment to the RECOVERY trial, and future studies, were included within the scope of the consensus process.

A Delphi process is a well-established method for achieving consensus from multiple groups of stakeholders(10), and has been used within healthcare for multiple reasons, including development of core outcome sets and identification of metrics for monitoring quality of care (11-16). Broadly, a Delphi process involves asking respondents to complete sequential questionnaires with group opinion fed-back to individual participants in between completion of the questionnaires. Children with PIMS-TS require the expertise of clinicians who specialise in immunology, infectious diseases, respiratory, rheumatology, cardiology, intensive care, general paediatrics, haematology and in some cases surgery, radiology and neurology. The aims of this study were therefore to seek consensus from participants within these key stakeholder groups regarding the diagnosis and management of children with suspected PIMS-TS, to identify areas where equipoise existed in order to inform subsequent research, and to explore whether consensus existed with regards to how children with PIMS-TS could be enrolled in the RECOVERY trial.

## Methods

### Ethics approval

This work was considered quality improvement by the Health Research Authority, and therefore approval by an ethics review board was not required.

### Summary

A three-phase online Delphi process was used to identify statements where a national multidisciplinary panel agreed that consensus existed regarding the investigation and management of children with suspected PIMS-TS. A consensus meeting was conducted via a web-based platform to review statements where consensus had not been achieved during the Delphi process. A face to face consensus meeting was not conducted due to COVID-19 social distancing restrictions.

### Scope

The consensus statements are applicable to children in the UK suspected of having PIMS-TS. They may also be applicable in other high-income countries, although the views described only represent those of UK clinicians. They are less likely to be applicable in countries where infrastructure and access to healthcare and treatments are significantly different to that of the UK.

### Participants

Clinicians were purposively selected to cover the range of multidisciplinary clinical and research expertise needed to diagnose and manage children with PIMS-TS, and were invited personally, by email or by telephone to participate in the study through sub-speciality groups and personal contacts. Those who agreed to participate were divided into three panels in order to facilitate feedback throughout the Delphi process:

1. Paediatric Infectious Diseases and Immunology, Paediatric Rheumatology, Paediatric Respiratory, Pharmacist with specialist expertise in biological therapy
2. Paediatric Cardiology, Paediatric Intensive Care and Transport, Paediatric Haematology
3. General Paediatricians, Paediatric Radiologists and Paediatric Surgeons.

Representation in all three panels was sought, but experience in management of children with PIMS-TS, and the need to rapidly conclude the consensus process were prioritised over seeking wider engagement of clinicians or achieving numerical balance between the panels.

### Information sources

Statements for assessment in phase one of the Delphi process were derived by the study management group from reviews of the existing literature and expert opinion, including draft local guidelines. Participants in the Delphi process were asked in phase one and phase two to propose additional statements which they considered necessary for assessment. These were reviewed by the study management group, and if falling within the scope of the study, were included for assessment in the subsequent phase.

### Consensus process

A three-phase online Delphi process was conducted concurrently for the three panels. Results of the Delphi process were discussed in a virtual, online, consensus meeting attended by a representative sample of experts from each panel. The consensus meeting was chaired by an independent, non-voting, non-paediatric clinical academic experienced in Delphi methodology.

In phase one of the Delphi process, participants were asked to score statements from 1-9 based on how much they agreed with the statement. Scores of 1, 2 and 3 were ‘disagree with statement’, 4, 5 and 6 were ‘agree with statement’ and 7, 8 and 9 were ‘strongly agree with statement. Participants were asked to score a statement ‘don’t know’ if they did not consider themselves to have expertise in that area. In phase two, participants were shown graphical and numerical representations of how their panel overall had scored each statement and were asked to re-score the statements taking that information into account. In phase three, participants were shown graphical and numerical representations of how all three panels had scored each statement and asked to re-score the statements taking that information into account.

Participants were sent a reminder email if they had not completed the phase with 24 hours remaining. Participants who did not complete a phase were deemed to have withdrawn from the study and were not invited to take part in subsequent phases.

### Consensus definitions

‘Consensus agreement’ was defined as ≥70% of participants scoring a statement 7-9, and <15% of participants scoring a statement 1-3 in all three individual panels. ‘Consensus disagreement’ was defined as ≥70% of participants scoring a statement 1-3, and <15% of participants scoring a statement 7-9 in all three individual panels. Following phases two and three, if statements met ‘consensus agreement’ or ‘consensus disagreement’, they were excluded from the next stage of assessment.

Statements where consensus had been achieved in two out of three panels at the end of phase three were discussed in the consensus meeting. Statements discussed at the consensus meeting were assessed using a simple binary vote of ‘agree’ or ‘disagree’. Those statements where more than 70% of participants either agreed or disagreed with the statement were deemed to have met consensus. If consensus was not met following the initial vote, in depth discussions were held to understand why disagreement existed and were followed up with a second vote. Where participants felt agreement could be achieved with minor modifications to the statements, these modifications were made.

### Formation of the guidance

The final guidance is formed from those statements which met ‘consensus agreement’ or ‘consensus disagreement’ after phase two, phase three, or the consensus meeting.

### Patient and public involvement

Whilst in most healthcare related Delphi processes it has been appropriate to involve patients or the public as key stakeholders, it was felt that the clinical expertise required to assess the statements around which consensus was required for development of this clinical management pathway precluded inclusion of these groups. Patients and the public were therefore not involved in either the design or conduct of this study.

## Results

A total of 98 participants were invited to contribute in phase one of the Delphi process, 46 (47%) of whom completed all three phases (Table 1). Nine participants attended the consensus meeting (Table 1). 217 statements were assessed in phase one, 35 statements were added for assessment in phase two, and 3 statements added for assessment in phase three. Following phase two, 68 statements met consensus agreement, and 14 statements met consensus disagreement, and therefore a total of 82 statements were dropped from phase three. Of the 173 statements assessed in phase three, 22 met consensus agreement, and 10 met consensus disagreement, and were therefore dropped from the consensus meeting. 102 statements achieved consensus in no panels or only one panel and were therefore excluded from discussion at the consensus meeting. 39 statements met consensus for agreement or disagreement in two out of three panels and were therefore eligible for discussion at the consensus meeting. Six of these statements, where ‘consensus disagreement’ had been achieved in two panels were not discussed, as their recommendations were already covered by statements where consensus agreement had been achieved, and they would have been superfluous in the final guidance. Of the statements discussed, 21 met consensus for agreement, and 5 met consensus for disagreement following the consensus meeting. At the end of the process, there was consensus agreement for 111 statements, consensus disagreement for 29 statements, and no consensus achieved for 115 statements. The final guidance is therefore formed from the 140 statements where consensus agreement was achieved throughout the consensus process (Figure 2). Supplementary material 1 lists all assessed statements and their final consensus decisions. Figure 3 and Boxes 1a – 4a summarise the guidance based upon the 140 statements where consensus was achieved.

**Table 1:**
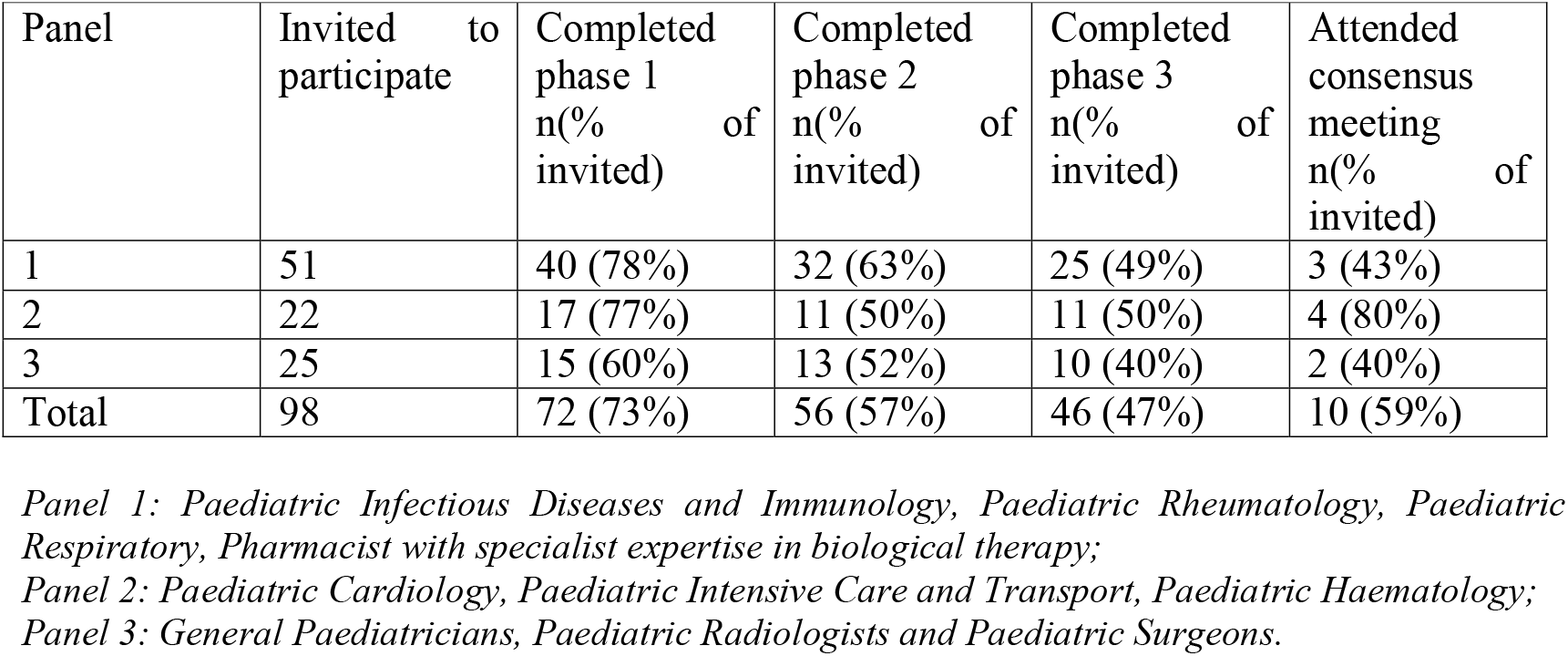
Participants.

| Panel | Invited to participate | Completed phase 1<br>n(% of invited) | Completed phase 2<br>n(% of invited) | Completed phase 3<br>n(% of invited) | Attended consensus meeting<br>n(% of invited) |
| --- | --- | --- | --- | --- | --- |
| 1 | 51 | 40 (78%) | 32 (63%) | 25 (49%) | 3 (43%) |
| 2 | 22 | 17 (77%) | 11 (50%) | 11 (50%) | 4 (80%) |
| 3 | 25 | 15 (60%) | 13 (52%) | 10 (40%) | 2 (40%) |
| Total | 98 | 72 (73%) | 56 (57%) | 46 (47%) | 10 (59%) |

*Panel 1: Paediatric Infectious Diseases and Immunology, Paediatric Rheumatology, Paediatric Respiratory, Pharmacist with specialist expertise in biological therapy;*
*Panel 2: Paediatric Cardiology, Paediatric Intensive Care and Transport, Paediatric Haematology;*
*Panel 3: General Paediatricians, Paediatric Radiologists and Paediatric Surgeons.*

**Fig 1.**
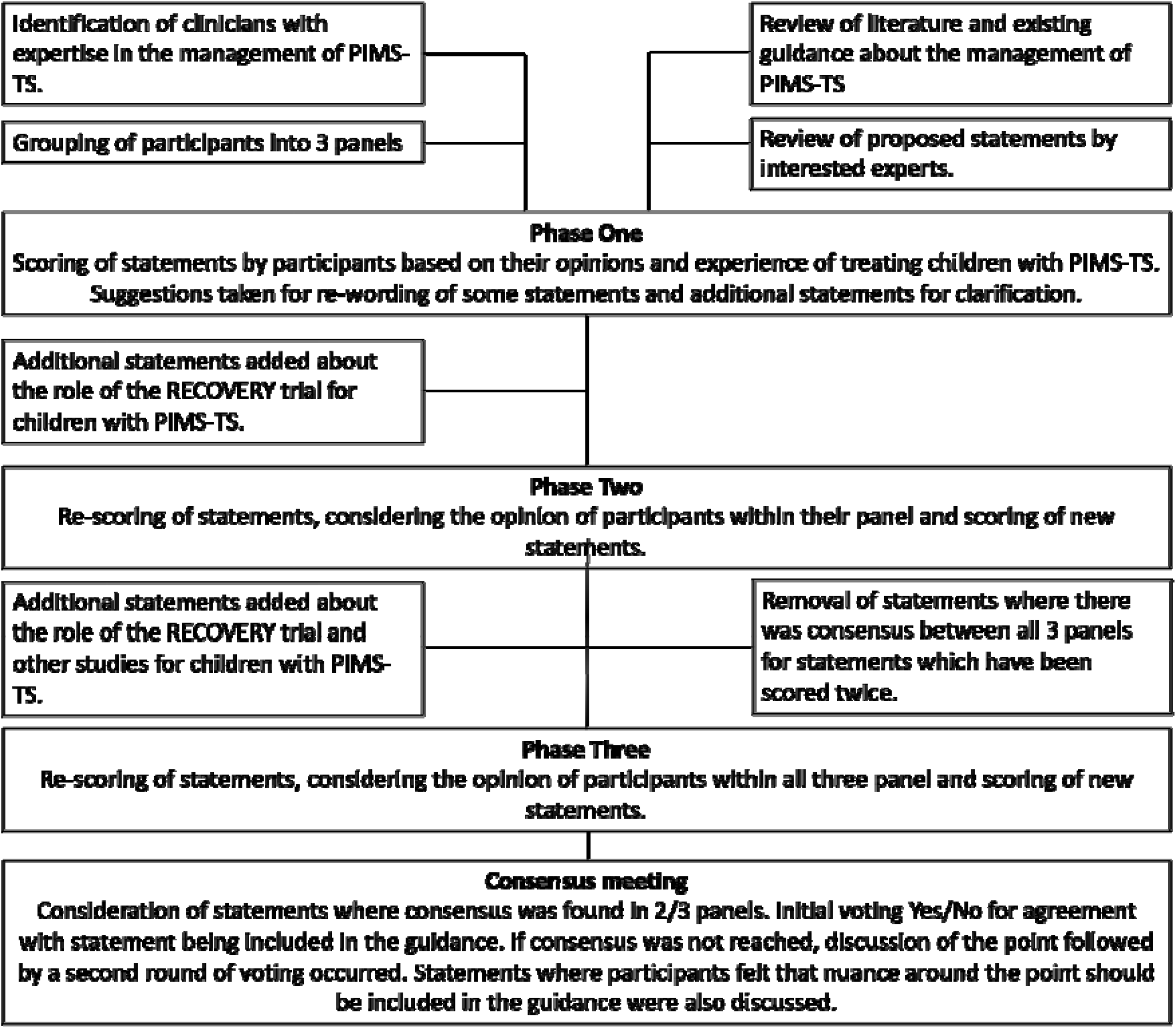

**Fig 2.**
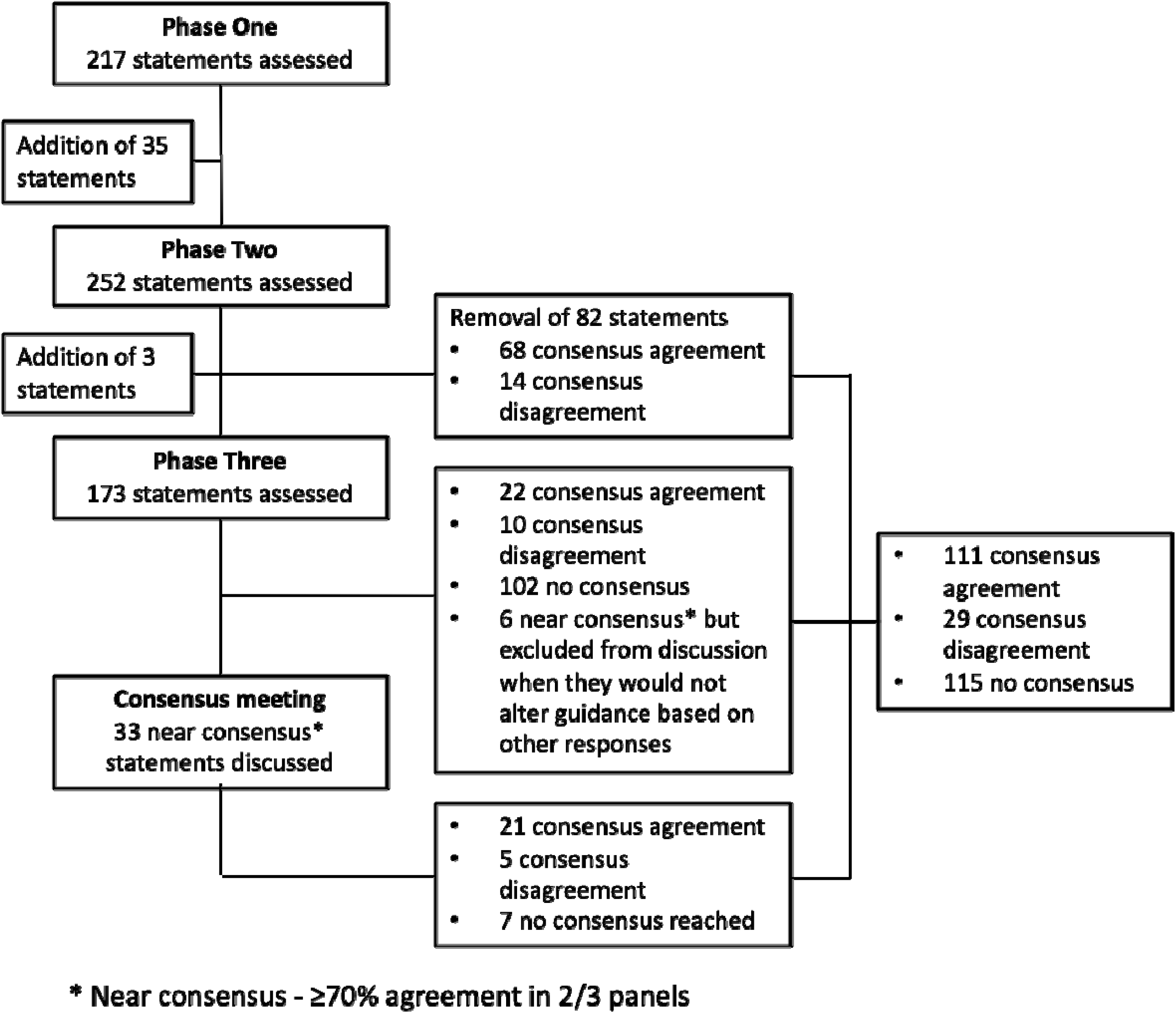

**Figure 3:**
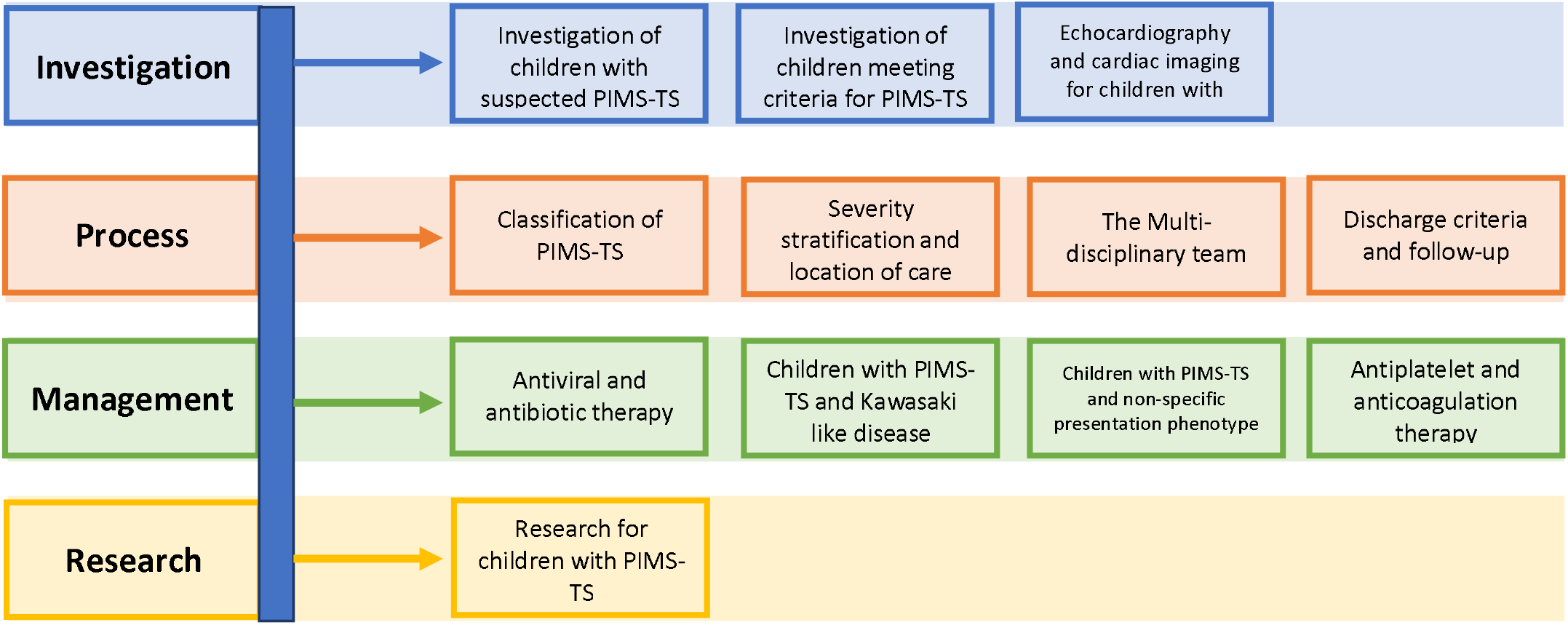
Areas covered by the PIMS-TS consensus guidance.

## Discussion

Use of an online Delphi process and virtual consensus meeting has enabled a National multidisciplinary panel to achieve consensus around 140 statements relating to the investigation and management of children with PIMS-TS, and participation of these children in studies including, but not limited to, DIAMONDS (https://www.diamonds2020.eu), ISARIC CCP-UK (https://isaric4c.net) and the RECOVERY trial (https://www.recoverytrial.net). Based upon the results of this process, it has been possible to develop a national consensus management pathway for the care of children with suspected PIMS-TS within 6 weeks of the need for such guidance becoming apparent. However, all participants recognise that this process has relied on clinical opinion based upon the limited evidence currently available. Until further evidence materialises, this management pathway can provide a framework for managing children with suspected PIMS-TS. The sections relating to research will help ensure the views of a wide range of clinicians are taken into account when seeking amendments for existing trials or designing future trials. Taking account of these views could maximise clinicians’ willingness to recruit to clinical trials.

The key strength of this work was the ability to achieve consensus relating to the management of a novel, complex condition, based upon quantitative data, from a relatively large number of participants, spread across multiple geographic regions. It was conducted in a short period of time, in the middle of a global pandemic, without the ability to conduct face to face meetings, large round table discussions, or focus groups. To our knowledge, such a process has not been attempted before. There are however three key limitations to the study. Firstly, the output and recommendations from a Delphi process can only ever be as robust as the statements that are assessed within it. As the statements assessed here were all developed based-upon level five evidence (expert opinion), the guidance can only ever seek to summarise this expert opinion. Once higher levels of evidence become available, these should be incorporated into future guidance in order to ensure that the management pathway remains relevant and up to date. Given the cost-efficient, timely nature of the conducted Delphi process, it would be feasible to re-run the process when significant new data comes to light, and to use the results of the process to inform development of guidance. The second limitation of the study is that a smaller number of participants were recruited from stakeholder groups than would normally be aimed for in conduct of a Delphi process, and the scope of the work precluded inclusion of parents, or members of the public in the process. Despite this, adequate representation was achieved across all panels, with multiple representatives from each stakeholder group participating. However, had time, and the need to ensure clinical expertise of participants not been such pressing factors, it would have been preferable to seek opinions from a larger number of stakeholders. Finally, the consensus meeting included only a few representatives of each stakeholder group due to the online format and need to ensure opinions from all stakeholder groups during the meeting.

The management pathway created from this consensus process generally aligns well with the small evidence base that currently exists for similar clinical conditions. In particular, the guidance focuses on the recognition of severe cardiac disease which has been described in both phenotypes of PIMS-TS(5, 17). It includes a management pathway for Kawasaki-like Disease which aligns with current guidance for the management of Kawasaki Disease(18), and may help to address the current variation in treatment which is occurring regarding the indications for intravenous immunoglobulin (IVIg)(17). Discussion and voting during the consensus meeting found equipoise for randomising within trials between IVIg and Methylprednisolone for both phenotypes of PIMS-TS. There was support within the consensus group that it would be appropriate for a trial to consider ‘supportive care only’ as an additional arm, but this was not voted on and therefore not included in the consensus guidance.

Within the Delphi process, significant discrepancy was noted between panel one (paediatric medical specialists with training in immunology) and panel two (paediatric intensivists, cardiologists and haematologists) with regards to whether children with PIMS-TS should be cared for in units with extra-corporeal membrane oxygenation (ECMO) availability. 90% of panel two strongly agreed this should be the case, whilst 86% of panel one disagreed. Data collected by the national British Paediatric Surveillance Unit (https://www.rcpch.ac.uk/work-we-do/bpsu) PIMS-TS surveillance study will help to provide the underpinning research to resolve this discrepancy. Until such data are available, we would reinforce the need for significant clinical decisions relating to the management of children with PIMS-TS to be taken within a multi-disciplinary setting, with adequate representation from all core members of the multidisciplinary team. Other areas where the need for future research have been highlighted by this Delphi process include identification of the most appropriate immunomodulatory therapy for use in children with the non-specific PIMS-TS phenotype, and whether IVIG or methylprednisolone should be first line therapy for children with both phenotypes of PIMS-TS.

This is the first published consensus management pathway relating to the treatment of children with PIMS-TS(8, 19). It is based on consensus expert opinion and is intended to act as a framework for the safe management of children with this condition. As new, higher level evidence become available, the guidance will be updated.

## Data Availability

Relevant raw data will be provided in supplementary material. No further data will be available for sharing.

## Competing Interests

All authors have completed the ICMJE uniform disclosure form at ww.icmje.org/coi_disclosure.pdf.

## Transparency statement

The lead author (the manuscript’s guarantor) affirms that the manuscript is an honest, accurate, and transparent account of the study being reported; that no important aspects of the study have been omitted; and that any discrepancies from the study as originally planned have been explained.

## Funding sources

NHS England funded the software support that was required to collect responses. Marian Knight and Saul Faust are NIHR Senior Investigators. Benjamin Allin is funded by an NIHR Doctoral Research Fellowship. Rachel Harwood holds a KRUK training fellowship. The views expressed are those of the author(s) and not necessarily those of the NHS, the NIHR or the Department of Health.

## Author Contributions

RH and BA designed and conducted the study with SK with additional input from SNF, CEJ, MK and RA. RH and BA wrote the paper with detailed input from SK, MK, SNF and CEJ. All authors were involved in study analysis and approved the final manuscript.

## Patient and Public Involvement

There was no involvement of patients or their families in this management pathway. We recognise their important role in the care of children with PIMS-TS and intend to consult them for future iterations of the pathway.

## Acknowledgements

We would like to thank Marcel Minke and Gabriel Jenik for their support in developing the customised Limesurvey database that made conduct of the Delphi process possible within the required timeframe. We would also like to thank Joseph Skelton whose organisational skills made this process possible and the Royal College of Paediatrics and Child Health, in particular Professor Russell Viner, Professor Nick Bishop and Carmel Turner, who gave their support to this project.

The PIMS-TS National Consensus Management Study Group consists of Rachel Agbeko, Octavio Aragon, Jim Baird, Alasdair Bamford, Michael Beresford, Tara Bharucha, Paul Brogan, Karina Butler, Enitan Carroll, Katrina Cathie, Ashish Chikermane, Sharon Christie, Matthew Clark, Antigoni Deri, Conor Doherty, Simon Drysdale, Phuoc Duong, Saravanan Durairaj, Marieke Emonts, Jennifer Evans, James Fraser, Scott Hackett, Rosie Hague, Paul Heath, Jethro Herberg, Marina Ilina, Nicola Jay, Dominic Kelly, Caroline Kerrison, Jeanette Kraft, Alice Leahy, Mike Linney, Hermione Lyall, Liza McCann, Paddy McMaster, Owen Miller, Sean O’Riordan, Stephen Owens, Clare Pain, Sanjay Patel, Nazima Pathan, Jim Pauling, David Porter, Andy Prendergast, Ravi Kumar, Andrew Riorden, Marion Roderick, Barney Schofield, Malcolm G Semple, Ethan Sen, Fiona Shackley, Ian Sinha, Shane Tibby, Andrew Tometzki, Stefania Verganano, Steven Welch, Nick Wilkinson, Mark Wood and Iain Yardley in addition to the authors of this manuscript.

## Guidance

**RCPCH Definition of Paediatric Inflammatory Multisystem Syndrome Temporally Associated with SARS-CoV-2 (PIMS-TS)**

The following set of statements refers to the Paediatric Inflammatory Multisystem Syndrome temporally associated with SARS-CoV-2 infection (PIMS-TS), defined by the Royal College of Paediatrics and Child Health:

A child presenting with persistent fever, inflammation (neutrophilia, elevated CRP and lymphopaenia) and evidence of single or multi-organ dysfunction (shock, cardiac, respiratory, renal, gastrointestinal or neurological disorder) with additional features. This may include children fulfilling full or incomplete criteria for Kawasaki Disease.

Exclusion of any other microbial cause, including bacterial sepsis, staphylococcal or streptococcal shock syndromes, infections associated with myocarditis such as enterovirus.

Positive or negative SARS-CoV-2 PCR

### Box 1a

**Investigation**

**Initial investigation of children with suspected PIMS**

1. Children presenting to hospital with fever, abdominal pain, gastro-intestinal, respiratory or neurological symptoms who are not unstable and have no other clear cause for their symptoms should have the following initial blood tests performed to help to identify whether they have PIMS-TS:
  a. Full blood count
  b. C-Reactive Protein
  c. Urea, creatinine and electrolytes
  d. Liver function tests

Footnote: The current definition for PIMS-TS includes persistent fever (>3 days) as a presenting complaint. As more cases are reported this may change but currently most experts feel that PIMS-TS should only be considered in febrile children. The ongoing study by the British Paediatric Surveillance Unit will provide further details around this.

### Box 1b

**Investigation**

**Haematological and biochemical investigation of children who meet the criteria for PIMS**

1. In addition to the tests above, children presenting with features which meet the criteria for PIMS-TS should have measurement of the following within 12 hours of admission:
  a. Blood gas and lactate
  b. Fibrinogen
  c. Ferritin
  d. D-Dimer
  e. Troponin
  f. N-terminal pro-B-type natriuretic peptide (NT-proBNP)*
  g. Lactate Dehydrogenase

* or equivalent

### Box 1c

**Investigation**

**Additional investigations for children who meet the criteria for PIMS**

1. Children presenting with features which meet the criteria for PIMS-TS should have the following investigations:
  a. SARS-CoV-2 reverse transcriptase polymerase chain reaction (RT-PCR) test on an appropriate respiratory sample ***and*** SARS-CoV-2 serology
  b. Septic and viral screen (lumbar puncture only if specifically indicated)
  c. 12 lead Electrocardiogram (ECG)
  d. Chest Radiograph
  e. Echocardiogram
2. In children with abdominal pain who meet the criteria for PIMS-TS and require imaging, abdominal ultrasound scan should be the first-line investigation.
3. Echocardiogram is not routinely recommended for children presenting with symptoms which *do not* meet the criteria for PIMS-TS.
4. Children who are physiologically unstable should have a daily echocardiogram
5. There is no consensus about the frequency of subsequent echocardiograms for physiologically stable children with PIMS-TS. We recommend that this is determined by a paediatric cardiologist based on the previous echocardiography findings, the clinical status of the patient and the change in blood markers of inflammation.
6. All children with coronary artery dilatation should be discussed with a paediatric cardiologist
7. Contrast enhanced computed tomography of the coronary vessels is not routinely recommended for children with PIMS-TS

### Box 2a

**Process**

**Classification of PIMS**

1. Primary classification of PIMS should be based on the presenting phenotype:
  a. Kawasaki-like Disease: Complete and incomplete, classified using the American Heart Association criteria.
  b. Non-specific: children presenting with shock and/or fever and symptoms which may include abdominal pain, gastrointestinal, respiratory or neurological symptoms which do not meet the criteria for Kawasaki Disease.

Subsequent classification of severity should be undertaken.

### Box 2b

**Process**

**Location of care and features of severity of PIMS**

1. Location of care should be determined by the severity of disease and MDT discussion will aid risk stratification for determining this.
2. Features of severe disease may be signified by the presence of any of the following factors, particularly when present in combination:
  - Physiological features of severe disease
    a. Prolonged capillary refill time
    b. Persistent hypotension
    c. Persistent tachycardia
    d. Requirement for 40ml/kg fluid bolus
    e. Oxygen saturations less than 92% in room air
  - Haematological and biochemical features
    a. Significantly elevated C-Reactive Protein (consensus reached for >300 mg/L but subsequent evidence suggests >150 mg/L)
    b. Significantly elevated or rising troponin
    c. Elevated Brain Natriuretic Peptide
    d. Elevated or rising lactate
    e. Elevated creatinine
    f. Significantly elevated or rising D-Dimer
    g. Elevated or rising Lactate Dehydrogenase
    h. High or low Fibrinogen
  - Cardiac features
    a. Abnormal ECG
    b. Coronary Artery Aneurysms on echocardiogram
    c. Left ventricular failure on echocardiogram
3. Children with features of complete or incomplete Kawasaki Disease-like phenotype can be cared for in a District General Hospital if (1) they do not have single or multiple organ dysfunction or cardiac disease (2) they can have an echo by a clinician with a competency to assess for cardiac involvement including coronary artery abnormalities.
4. Children with any evidence of cardiac involvement (elevated Troponin, elevated BNP, abnormal coronary arteries on echo or contrast enhance computed tomography) should be cared for in a level 2 or level 3 unit where there is specialist cardiology cover on-site.
5. Any child with single or multiple organ dysfunction should be cared for in a level 3 unit with specialist cardiology cover on-site.

### Box 2c

**Process**

**Multi-Disciplinary Team**

1. Early discussion, before formal MDT, should occur for severely unwell children.
2. Every child with PIMS-TS should be discussed by an MDT within 24 hours of admission or identification of PIMS-TS if already an inpatient.
3. Core members of the multi-disciplinary team include:
  a. Paediatric Infectious Diseases/Immunologists
  b. AND/OR Paediatric Rheumatologists
  c. AND Paediatric Cardiologists
  d. AND Paediatric Intensivists
4. Additional members of the multi-disciplinary team include:
  a. General paediatricians caring for children in a District General Hospital and for children with multiple co-morbidities
  b. Paediatric Emergency Transport Team - for children who are severely unwell in a District General Hospital at the time of MDT
  c. Paediatric Haematologists - for children with haemaglobinopathies, clotting disorders, coagulopathy or thrombosis

### Box 2d

**Process**

**Discharge criteria and follow-up**

1. To be discharged from hospital, children who are otherwise well should have
  a. Stable cardiac function
  b. No pyrexia for 24 hours
2. Children with PIMS should be followed up in the first 1-2 weeks after discharge and have further follow-up 6 weeks after discharge. Echocardiography should form part of this follow-up.

**Multi-disciplinary follow-up**

1. Multi-disciplinary follow-up should be undertaken for children:
  a. with coronary artery abnormalities
  b. who have required organ support due to PIMS
2. Multi-disciplinary clinicians should include:
  a. Paediatric Cardiology
  b. Paediatric Infectious Diseases

### Box 3a

**Management**

**Anti-viral and antibiotic therapy**

1. Children with PIMS who are SARS-CoV-2 positive on rt-PCR or antigen testing, may be considered for anti-viral therapy. Remdesivir is the first-choice anti-viral therapy.
2. IV antibiotics should be commenced in all patients. These should be focussed or stopped based on the clinical picture and culture results.
3. Children who meet the criteria for toxic shock syndrome should receive clindamycin in addition to broad spectrum antibiotics.
4. The initial infection screen does not have to be negative for other pathogens prior to high dose steroids being commenced.

### Box 3b

**Management**

**Management of children with PIMS and features of Kawasaki-like Disease (Complete and incomplete phenotype)**

1. First line therapy for all children is IVIg at a dose of 2g/kg, calculated using ideal body mass index. This can be administered in a single or divided dose depending on the clinical picture and cardiac function.
  a. A second dose of IVIg may be considered for children who have not responded or partially responded to the first dose of IVIg.
2. All children who meet the criteria for the RECOVERY trial should be invited to participate in the first stage randomisation.
3. ‘High risk’ children include those under 12 months of age and those with coronary artery changes. These children should receive early IV methylprednisolone (i.e. alongside IVIg)*.
4. If a child is recruited to the first randomisation in the RECOVERY trial they will be randomised between therapy and ‘standard of care’. At present, the RECOVERY first randomisation would be in addition to the standard of care described below.
5. Second line therapy is IV Methylprednisolone*. It should be considered as the next treatment option for children who remain unwell 24 hours after IVIg infusion, particularly if they have ongoing pyrexia.
6. Gastric Protection should be given to children on high dose steroids.
7. Biological therapy should be considered as third line in children who fail to respond to IVIg and IV Methylprednisolone.
  a. The decision to commence a biological agent should be made by a multi-disciplinary team
  b. If a child is recruited to the RECOVERY trial they should be offered the opportunity to enter the 2^nd^ stage interventions phase and be randomised between Tocilizumab and standard of care. Stage 1 and 2 randomisations in the RECOVERY trial can happen at the same time.
  c. The preferred biological agent for children with Kawasaki-like Disease phenotype is infliximab.

*Please note: lower dose corticosteroid can be considered as a treatment option in the first randomisation to the RECOVERY trial, even if methylprednisolone has already been given.

### Box 3c

**Management**

**Management of children with PIMS and non-specific presentation phenotype**

1. First line therapy for all children is IVIg at a dose of 2g/kg, calculated using ideal body mass index. This can be administered in a single or divided dose depending on the clinical picture and cardiac function.
  a. A second dose of IVIg may be considered for children who have not responded or partially responded to the first dose of IVIg.
2. Indications for therapy include:
  a. Evidence of coronary artery abnormality
  b. Meeting the criteria for toxic shock syndrome
  c. Evidence of progressive disease
  d. Prolonged fever for >5 days
3. All children who meet the criteria for the RECOVERY trial should be invited to participate in the first stage randomisation.
4. If a child is recruited to the first randomisation in the RECOVERY trial they will be randomised between therapy and ‘standard of care’. At present, the RECOVERY first randomisation would be in addition to the standard of care described below.
5. Second line therapy is IV Methylprednisolone. It should be considered as the next treatment option for children who remain unwell 24 hours after IVIg infusion, particularly if they have ongoing pyrexia.*
6. Gastric Protection should be given to children on high dose steroids.
7. Third line therapy is a biological agent in children who fail to respond to IVIg and IV Methylprednisolone.
  a. The decision to commence a biological agent should be made by a multi-disciplinary team
  b. If a child is recruited to the RECOVERY trial they should be offered the opportunity to enter the 2^nd^ stage interventions phase and be randomised between Tocilizumab and standard of care. Stage 1 and 2 randomisations in the RECOVERY trial can happen at the same time.
  c. Consensus was not reached on a preferred biological agent with equipoise demonstrated between Tocilizumab, Anakinra and Infliximab.
8. A small number of children within this phenotype have met the criteria for haemophagocytic lymphohistiocytosis (HLH). In these children discussion with a specialist team and awareness of the HLH-2004 guidelines is recommended.

*Please note: lower dose corticosteroid can be considered as a treatment option in the first randomisation to the RECOVERY trial, even if methylprednisolone has already been given.

### Box 3d

**Management**

**Anti-platelet and anti-coagulation therapy for children with PIMS**

1. All children over 12 years of age should wear compression stockings.
2. The local Kawasaki guideline for aspirin dosing should be followed for children with Kawasaki-like Disease Phenotype.
3. No consensus was reached over whether children with non-specific phenotype should receive high dose aspirin in specific situations.
4. Low dose aspirin should be continued for a minimum of 6 weeks in all patients with PIMS-TS.
5. Children who have a thrombotic event should follow the local protocol for management of this.
6. Children with abnormal coronary arteries should be discussed with a haematologist regarding long-term anti-platelet therapy and anti-coagulation.

### Box 4

**Research for children with PIMS-TS**

**RECOVERY trial**

RECOVERY is an adaptive trial and based on this Delphi process, the Trial Steering Committee are considering a PIMS-TS specific first randomisation protocol amendment.

1. All children who meet the criteria for inclusion in the RECOVERY trial should be offered the opportunity to enter and be randomised in the first stage.
2. For a future amendment in RECOVERY or a future research trial there is equipoise for children with both phenotypes of PIMS to receive methylprednisolone OR IVIg as a first line treatment within a research study.
3. Children enrolled in the RECOVERY trial should be offered the opportunity to enter the 2nd stage intervetions arm (Tocilizumab vs standard of care) if they have been discussed by a MDT and the decision made to commence biological therapy.
4. For a future research trial there is equipoise between tocilizumab, anakinra and infliximab for patients with a non-specific phenotype.
5. Infliximab is the preferred biologic for children with Kawasaki-like Disease.
6. All families should be approached for inclusion in research studies including DiAMONDS and ISARIC/CCP-UK. All children should be enrolled to the national BPSU PIMS-TS registry.

### Box 5

**References and resources for guidance**

1. BW McCrindle et al. Diagnosis, Treatment and Long-Term Management of Kawasaki Disease: A scientific statement for Health Professionals From the American Heart Association. Circulation. 2017;135(17)
2. J-I Henter et al, HLH-2004: Diagnostic and Therapeutic Guidelines for Haemophagocytic Lymphohistiocytosis. Pediatr Blood Cancer, 2007;48(2)
3. Diamonds: https://imperialbrc.nihr.ac.uk/research/covid-19/covid-19-ongoing-studies/diamonds/
4. ISARIC-CCP: https://isaric.tghn.org/UK-CCP/
5. RECOVERY: https://www.recoverytrial.net/

## Notes

### Competing Interest Statement

The authors have declared no competing interest.

## References

1. Jones VG, Mills M, Suarez D, Hogan CA, Yeh D, Segal JB, et al. COVID-19 and Kawasaki Disease: Novel Virus and Novel Case. Hosp Pediatr. 2020;10(6):537–40.

2. Toubiana J, Poirault C, Corsia A, Bajolle F, Fourgeaud J, Angoulvant F, et al. Outbreak of Kawasaki disease in children during the COVID-19 pandemic: a prospective observational study in Paris, France. medRxiv. 2020.

3. Belhadjer Z, Meot M, Bajolle F, Khraiche D, Legendre A, Abakka S, et al. Acute heart failure in multisystem inflammatory syndrome in children (MIS-C) in the context of global SARS-CoV-2 pandemic. Circulation. 2020.

4. Verdoni L, Mazza A, Gervasoni A, Martelli L, Ruggeri M, Ciuffreda M, et al. An outbreak of severe Kawasaki-like disease at the Italian epicentre of the SARS-CoV-2 epidemic: an observational cohort study. The Lancet. 2020;395(10239):1771–8.

5. Whittaker E, Bamford A, Kenny J, Kaforou M, Jones CE, Shah P, et al. Clinical Characteristics of 58 Children With a Pediatric Inflammatory Multisystem Syndrome Temporally Associated With SARS-CoV-2. JAMA. 2020.

6. Riphagen S, Gomez X, Gonzalez-Martinez C, Wilkinson N, Theocharis P. Hyperinflammatory shock in children during COVID-19 pandemic. The Lancet. 2020;395(10237):1607–8.

7. RCPCH. Guidance: Paediatric multisystem inflammatory syndrome temporally associated with COVID-19 2020. Available from: https://www.rcpch.ac.uk/sites/default/files/2020-05/COVID-19-Paediatric-multisystem-%20inflammatory%20syndrome-20200501.pdf.

8. CDC. Multisystem Inflammatory Syndrome in Children (MIS-C) Associated with Coronavirus Disease 2019 (COVID-19) 2020. Available from: https://emergency.cdc.gov/han/2020/han00432.asp.

9. WHO. Multisystem inflammatory syndrome in children and adolescents with COVID-192020. Available from: https://apps.who.int/iris/handle/10665/332095.

10. Okoli C, Pawlowski SD. The Delphi method as a research tool: an example, design considerations and applications. Information & Management. 2004;42(1):15–29.

11. Sinha IP, Smyth RL, Williamson PR. Using the Delphi technique to determine which outcomes to measure in clinical trials: recommendations for the future based on a systematic review of existing studies. PLoS Med. 2011;8(1):e1000393.

12. Dalkey N, Helmer O. An Experimental Application of the DELPHI Method to the Use of Experts. Management Science. 1963;9(3):458–67.

13. Bunch KJ, Allin B, Jolly M, Hardie T, Knight M. Developing a set of consensus indicators to support maternity service quality improvement: using Core Outcome Set methodology including a Delphi process. Bjog. 2018;125(12):1612–8.

14. Allin BSR, Bradnock T, Kenny S, Kurinczuk JJ, Walker G, Knight M, et al. NETS(1HD) study: development of a Hirschsprung’s disease core outcome set. Arch Dis Child. 2017;102(12):1143–51.

15. Bamber JH, Lucas DN, Plaat F, Allin B, Knight M, Quality cftOAA, et al. The identification of key indicators to drive quality improvement in obstetric anaesthesia: results of the Obstetric Anaesthetists’ Association/National Perinatal Epidemiology Unit collaborative Delphi project. Anaesthesia. n/a(n/a).

16. Allin BSR, Hall NJ, Ross AR, Marven SS, Kurinczuk JJ, Knight M, et al. Development of a gastroschisis core outcome set. Arch Dis Child Fetal Neonatal Ed. 2019;104(1):F76–F82.

17. Toubiana J, Poirault C, Corsia A, Bajolle F, Fourgeaud J, Angoulvant F, et al. Kawasaki-like multisystem inflammatory syndrome in children during the covid-19 pandemic in Paris, France: prospective observational study. BMJ. 2020;369:m2094.

18. McCrindle BW, Rowley AH, Newburger JW, Burns JC, Bolger AF, Gewitz M, et al. Diagnosis, Treatment, and Long-Term Management of Kawasaki Disease: A Scientific Statement for Health Professionals From the American Heart Association. Circulation. 2017;135(17):e927–e99.

19. Control) EECfDPa. Paediatric inflammatory multisystem syndrome and SARS-CoV-2 infection in children 2020.

